# SARS-CoV-2 antigens expressed in plants detect antibody responses in COVID-19 patients

**DOI:** 10.1101/2020.08.04.20167940

**Authors:** Mohau S. Makatsa, Marius B. Tincho, Jerome M. Wendoh, Sherazaan D. Ismail, Rofhiwa Nesamari, Francisco Pera, Scott de Beer, Anura David, Sarika Jugwanth, Maemu P. Gededzha, Nakampe Mampeule, Ian Sanne, Wendy Stevens, Lesley Scott, Jonathan Blackburn, Elizabeth S. Mayne, Roanne S. Keeton, Wendy A. Burgers

**Affiliations:** Institute of Infectious Disease and Molecular Medicine, University of Cape Town, Cape Town, South Africa; Division of Medical Virology, Department of Pathology, University of Cape Town, Cape Town, South Africa; Cape Bio Pharms, Cape Town, South Africa; Department of Molecular Medicine and Haematology, University of Witwatersrand, Johannesburg, South Africa; Department of Immunology, Faculty of Health Sciences, University of Witwatersrand and National Health Laboratory Service Johannesburg, South Africa; Clinical HIV Research Unit, Department of Internal Medicine, University of Witwatersrand, Johannesburg, South Africa; Division of Chemical and Systems Biology, Department of Integrative Biomedical Sciences, University of Cape Town, Cape Town, South Africa; Wellcome Centre for Infectious Diseases Research in Africa, University of Cape Town, Cape Town, South Africa

**Author notes:** **Corresponding author:** Wendy Burgers, Institute of Infectious Disease and Molecular Medicine, Faculty of Health Sciences, University of Cape Town, Observatory 7925, South Africa. M.S.M., M.B.T. and J.M.W. contributed equally to this work. **Declaration of interests:** Francisco Pera and Scott de Beer are employed by Cape Bio Pharms. The remaining authors declare that the research was conducted in the absence of any commercial or financial relationships that could be construed as a potential conflict of interest. **Author contributions:** Conceived and designed the study and experiments: WAB, MSM, MBT, JMW, FP, SdB, ESM, LS and JB. Provided support and critical protocol review: WS, IS. Performed the experiments: MSM, MBT, JMW, AD, SJ, NM, MG, FP, SdB. Analyzed the data: MSM, AD, SJ, NM, MG, SDI, RN, RSK and WAB. Wrote the paper: FP, SDI, RN, RSK, MBT, JMW, MSM and WAB. All authors approved the final manuscript. **Contribution to the Field Statement** The SARS-CoV-2 pandemic poses a significant global threat to lives and livelihoods, with over 16 million confirmed cases and at least 650 000 deaths from COVID-19 in the first 7 months of the pandemic. Developing tools to measure antibody responses and understand protective immunity to SARS-CoV-2 is a priority. Many expression systems exist to produce the proteins required in the establishment of these serological assays, but plant-based systems have several advantages over more widely used conventional protein expression systems. Most notably, they are rapid, scaleable and cost-effective, making them attractive protein expression systems particularly in low-income settings such as ours in Africa. We were able to develop a cost-effective serological assay by making use of plant-produced viral antigens. Our study demonstrates that recombinant SARS-CoV-2 proteins produced in plants enable the robust detection of SARS-CoV-2-specific antibodies equivalent to that observed in a high sensitivity commercial assay in which antigens were produced in a mammalian expression system. Our ELISA can be used to evaluate SARS-CoV-2 seroprevalence, describe the kinetics of the humoral immune response in infected individuals, and investigate humoral immunity in our setting where comorbidities are highly prevalent.

**Keywords:** SARS-CoV-2, COVID-19, serology, ELISA, plant expression

## Abstract

**Background:** The SARS-CoV-2 pandemic has swept the world and poses a significant global threat to lives and livelihoods, with over 16 million confirmed cases and at least 650 000 deaths from COVID-19 in the first 7 months of the pandemic. Developing tools to measure seroprevalence and understand protective immunity to SARS-CoV-2 is a priority. We aimed to develop a serological assay using plant-derived recombinant viral proteins, which represent important tools in less-resourced settings.

**Methods:** We established an indirect enzyme-linked immunosorbent assay (ELISA) using the S1 and receptor-binding domain (RBD) portions of the spike protein from SARS-CoV-2, expressed in *Nicotiana benthamiana*. We measured antibody responses in sera from South African patients (n=77) who had tested positive by PCR for SARS-CoV-2. Samples were taken a median of six weeks after the diagnosis, and the majority of participants had mild and moderate COVID-19 disease. In addition, we tested the reactivity of pre-pandemic plasma (n=58) and compared the performance of our in-house ELISA with a commercial assay. We also determined whether our assay could detect SARS-CoV-2-specific IgG and IgA in saliva.

**Results:** We demonstrate that SARS-CoV-2-specific immunoglobulins are readily detectable using recombinant plant-derived viral proteins, in patients who tested positive for SARS-CoV-2 by PCR. Reactivity to S1 and RBD was detected in 51 (66%) and 48 (62%) of participants, respectively. Notably, we detected 100% of samples identified as having S1-specific antibodies by a validated, high sensitivity commercial ELISA, and OD values were strongly and significantly correlated between the two assays. For the pre-pandemic plasma, 1/58 (1.7%) of samples were positive, indicating a high specificity for SARS-CoV-2 in our ELISA. SARS-CoV-2-specific IgG correlated significantly with IgA and IgM responses. Endpoint titers of S1- and RBD-specific immunoglobulins ranged from 1:50 to 1:3200. S1-specific IgG and IgA were found in saliva samples from convalescent volunteers.

**Conclusions:** We demonstrate that recombinant SARS-CoV-2 proteins produced in plants enable robust detection of SARS-CoV-2 humoral responses. This assay can be used for seroepidemiological studies and to measure the strength and durability of antibody responses to SARS-CoV-2 in infected patients in our setting.

## Introduction

The current global pandemic, caused by the novel severe acute respiratory syndrome coronavirus 2 (SARS-CoV-2), has resulted in over 16 million cases and at least 650 000 deaths, as of 27 July 2020. SARS-CoV-2 was first detected in December 2019 in Wuhan, a city in the Hubei province of China, and is thought to originate from zoonotic transmission of a bat coronavirus (Tan et al., 2020; Zhu et al., 2020). Coronavirus disease 2019 (COVID-19), the resultant disease, is commonly associated with fever, cough and fatigue, and in severe cases, pneumonia and respiratory failure (Chan et al., 2020).

SARS-CoV-2 is a 30kB positive-stranded RNA virus that is a member of the *Betacoronavirus* genus and the subgenus *Sarbecovirus* (Letko et al., 2020). The genus harbours human pathogens that cause respiratory infections, namely the highly virulent SARS-CoV and Middle East respiratory syndrome coronavirus (MERS-CoV), as well as the circulating ‘common cold’ human coronavirus (hCoV)-OC43 and hCoV-HKU1 (Su et al., 2016). Betacoronaviruses express four essential structural proteins, namely the spike (S) glycoprotein, membrane (M) protein, envelope (E) protein and nucleocapsid (N) protein, as well as multiple accessory and non-structural proteins (Neuman et al., 2011, Lu et al., 2020). The S glycoprotein is a homotrimer that protrudes from the surface of the viral particles (Tortorici and Veesler, 2019), and interacts with the human cell receptor angiotensin converting enzyme 2 (ACE2) through the receptor-binding domain (RBD), gaining viral entry into the host cell (Li 2016; Letko et al., 2020; Walls et al., 2020). S is cleaved by host cell proteases into two subunits: the S1 subunit which harbours the RBD and enables binding to host cell receptors, and the S2 subunit that is important for fusion with the host cell membrane (Walls et al., 2020; Wrapp et al., 2020).

The S1 subunit is highly immunogenic, and its RBD portion is the main target of neutralizing antibodies, thus becoming the focus of serological studies (Amanat et al., 2020; Huang et al., 2020; Liu et al., 2020; Okba et al., 2020). Recently, potent neutralizing antibodies isolated from the convalescent sera of SARS-CoV-2 patients were demonstrated to be protective against disease from high-dose SARS-CoV-2 challenge in a small animal model (Rogers et al., 2020), suggesting the potential for therapeutic interventions as well as inferring that recovered SARS-CoV-2 patients may be afforded protection from re-infection by neutralizing antibody responses. Amanat et al (2020) showed a strong correlation between the neutralizing antibody response and ELISA endpoint titers against S, suggesting the use of serological assays in estimating the percentage of infected people who have neutralizing antibodies that protect them from re-infection or disease.

Serological assays that can detect antibody responses to SARS-CoV-2 are critical for answering pressing questions regarding immunity to the virus. It is not known what proportion of infected individuals elicit antibodies to SARS-CoV-2, if antibodies serve as correlates of protection, and if so, what the threshold of binding or neutralizing titers are that will provide immunity, and the duration of these responses. Serological assays such as enzyme-linked immunosorbent assays (ELISA) can assist in answering these questions. These assays need to be both sensitive as well as demonstrate high specificity for SARS-CoV-2, and not give false positives due to cross-reactivity with widely circulating hCoVs NL63, 229E, OC43, and HKU1. While the N protein is more conserved among coronaviruses, the S protein sequence has lower sequence conservation. The S1 portion is 21-25% identical at the amino acid level to circulating hCoVs (Okba et al., 2020). Thus, serological assays using the full-length S protein, S1 subunit or RBD portion as antigens have shown good specificity with little cross-reactivity to NL63 and 229E (Amanat et al., 2020; Zhao et al., 2020) compared to the use of N protein (Zhao et al., 2020).

Purified recombinant proteins are essential for the establishment of serological assays. Numerous protein expression systems exist, each with their own advantages and limitations. These include bacterial, mammalian, yeast, insect and plant-based systems (Shanmugaraj et al., 2020, Yin et al., 2007). Plant-based systems have several advantages over more widely used conventional protein expression systems (Shanmugaraj and Ramalingam, 2014). Most notably, they are rapid, cost-effective and support post-translational modifications similar to mammalian cell systems, making them attractive protein expression systems particularly in low-income settings (Shanmugaraj and Ramalingam, 2014 and 2020; Maliga et al., 2004). Historically, their major disadvantage was low yield (Shanmugaraj et al., 2020), however advances in plant technology, including transient expression systems and viral vectors, have led to improvements in protein yield (Kapila et al., 1997; Shanmugaraj and Ramalingam, 2014; Yamamoto et al., 2018). Additionally, SARS-CoV S1 protein expressed in tomato and tobacco plants demonstrated good immunogenicity in mice (Pogrebnyak et al., 2005). Together, these studies highlight the potential of plant-based expression systems for the development of serological assay reagents as well as vaccines for the current SARS-CoV-2 pandemic.

In this study, we describe the development of an ELISA that enables detection of antibodies directed at the S1 subunit and the RBD portion of the SARS-CoV-2 S glycoprotein, generated through a plant-based expression system.

## Materials and methods

### Recombinant protein cloning and expression

The S1 portion and receptor binding domain (RBD) of the spike protein of SARS-CoV-2 Wuhan-Hu-1 isolate (GenBank: MN908947.3) were produced by Cape Bio Pharms, Cape Town, South Africa. Briefly, *Nicotiana benthamiana* codon-optimized DNA encoding S1 and RBD was synthesized commercially (Genscript). Both genes were fused at their C-terminal region to the fragment crystallizable region (Fc) of rabbit IgG1 (Genbank: L29172.1) and subsequently cloned into Cape Bio Pharms’ proprietary vector, pCBP2. *Agrobacterium tumefaciens* strain GV3101 (pMP90RK) was used to carry agroinfiltration. Growth of recombinant *A. tumefaciens* and vacuum infiltration of *N. benthamiana* plants was performed as described previously (Maclean et al., 2007). Three days post-infiltration, leaves were homogenized in the presence of phosphate buffered saline (PBS) at a 2:1 ratio buffer:leaf material. Cell debris was removed by centrifugation at 10 000 *g* for 10 min at 4°C, and the clarified supernatant was used for expression analyses and purification by Protein A affinity chromatography.

For purification, the extract was filtered through a 0.22 µm cellulose nitrate filter (Sartorius) before loading onto a pre-equilibrated 5 ml column packed with POROS MabCapture A resin (Thermo Fisher). The column was then washed with 10 column volumes of wash buffer (PBS, pH 7.5) and bound proteins eluted using elution buffer (0.1 M glycine, pH 2.5). Eluted fractions were captured in 1/10^th^ volume of neutralization buffer (1 M Tris, pH 8.5) and then pooled and applied to a 10K molecular weight cutoff (MWCO) Amicon centrifuge tube (Millipore) for buffer exchange against PBS and sample concentration.

Mouse anti-rabbit IgG (γ-chain specific) horseradish peroxidase conjugate (1:2500; IgG-HRP, Sigma) was used in a standard SDS-PAGE and western blot analysis to examine purity of the recombinant proteins.

### Volunteer recruitment and sample collection

Samples were collected from SARS-CoV-2 infected volunteers (n=77) recruited from Gauteng and the Western Cape provinces of South Africa from 10 April 2020 to 26 May 2020. Volunteers had previously undergone a reverse transcriptase polymerase chain reaction (RT-PCR) test for SARS-CoV-2 from an upper respiratory tract (nose/throat) swab collected into viral transport media. Swabs were processed through approved assays in accredited public and private clinical laboratories. Inclusion criteria were age >=18 years and a confirmed positive PCR for SARS-CoV-2 on the national database of the National Health Laboratory Services (NHLS). Of the 77 participants, 34 (44%) had a second positive PCR result recorded within a week after the first positive test. With respect to disease severity, five participants were asymptomatic, 23 had mild disease (characterised by mild upper respiratory tract symptoms), 38 had moderate disease (defined by gastrointestinal symptoms or lower respiratory tract symptoms), and two had severe disease (admission to hospital). Serum and saliva samples were collected between 8 and 70 days after the first positive PCR test. Ethical approval for these studies was obtained from the Human Research Ethics Committee (HREC) of the University of Witwatersrand (M200468) and the University of Cape Town (UCT; 210/2020). All participants provided written, informed consent.

Pre-pandemic plasma (n=58) was obtained from banked human samples that were collected from participants recruited from Cape Town, South Africa in 2011-2012, from a study protocol approved by the HREC of the University of Cape Town (158/2010). Storage consent was provided by all participants, and approval for use of the samples in this study was obtained from the HREC, UCT. Samples came from participants who were HIV-infected (n=27) or HIV-uninfected (n=31). All participants had tested positive for exposure to *Mycobacterium tuberculosis* based on a positive IFN-γ-release assay (QuantiFERON-TB Gold In-Tube), *i*.*e*. were classified as having latent tuberculosis infection. The median age was 26 years (interquartile range [IQR]: 22-34 years) and 44/58 (76%) were female. All HIV-infected individuals were antiretroviral treatment (ART)-naive, with a median CD4 count of 591 cells/mm^3^ (IQR: 511-749).

All samples were treated with 1% Triton-X100 (Sigma) for 60 min at room temperature to inactivate any potentially live virus in the samples (Remy et al., 2019).

### Enzyme-linked Immunosorbent Assay (ELISA)

The ELISA protocol was adapted from a published protocol (Stadlbauer et al., 2020). Briefly, 96-well plates (Nunc MaxiSorp, Thermo Fisher) were coated at 4°C overnight with 50 µl of varying concentrations (1-4 µg/ml) of purified recombinant RBD or S1 proteins in PBS or bicarbonate buffer (both Sigma). The following day, plates were washed five times using an automated plate washer and incubated at room temperature in blocking solution (1% casein or 3% non-fat powder milk prepared in PBS with 0.1% Tween 20 (PBS-T)). After 1 h, the blocking solution was discarded and 100 µl of serum, plasma or saliva samples (at 1:50 dilution for sera/plasma and 1:10 for saliva) were added for 2 h at room temperature. Next, plates were washed five times and incubated with goat anti-human IgG (Fc-specific) peroxidase conjugate (1:5000; IgG-HRP, Sigma), or goat anti-human IgA (⍰-chain specific), F(ab’)_2_ fragment peroxidase conjugate (1:5000; IgA-HRP, Sigma) or goat anti-human IgM peroxidase conjugate (1:2000; IgM-HRP, Southern Biotech) for 1 h at room temperature. The plate was then developed using 100 µl O-phenylenediamine dihydrochloride (OPD; Sigma) for 12 min before the reaction was stopped with 50 µl 3M hydrochloric acid (HCl, Sigma). The plates were read at 490 nm using a Versamax microplate reader (Molecular Devices) using SoftMax Pro software (version 5.3). A cutoff for positivity was set at two standard deviations (SD) above the mean optical density (OD) of pre-pandemic samples. For determining endpoint titers, 2-fold serial dilutions were performed for 20 PCR+ samples and 40 pre-pandemic controls. Area under the curve (AUC) was determined and the positivity threshold was calculated as before, mean+2SD. All patient samples were also analysed using the anti-SARS-CoV-2 ELISA (IgG; Euroimmun), which uses the S1 domain of the spike protein, according to the manufacturer’s instructions.

### Statistical analysis

Statistical analyses were performed in Prism (GraphPad, version 8). Nonparametric tests were used for all comparisons. The Friedman test with Dunn’s multiple comparison test was used for matched comparisons; the Mann–Whitney U unmatched and Wilcoxon matched pairs t-tests were used for unmatched and paired samples, respectively. Spearman Rank tests were used for all correlations. AUC was calculated in Prism. A p value of <0.05 was considered statistically significant.

## Results

### SARS-CoV-2 antigen expression in plants

The S1 and RBD portions of the Spike protein of SARS-CoV-2 were expressed in *Nicotiana benthamiana* as fusions to the rabbit IgG Fc tag. Western blot and SDS-PAGE analysis revealed expression of purified S1 (**Figure 1A & B**) and RBD (**Figure 1C & D**) at the expected protein sizes of ∼140kDa and ∼100kDa, respectively. Higher molecular weight bands of ∼280kDa and ∼200kDa indicated possible dimer formation of S1 and RBD, respectively. In addition, lower molecular weight bands indicated potentially multiple cleavage products of S1 and RBD in the preparations.

**Figure 1.**
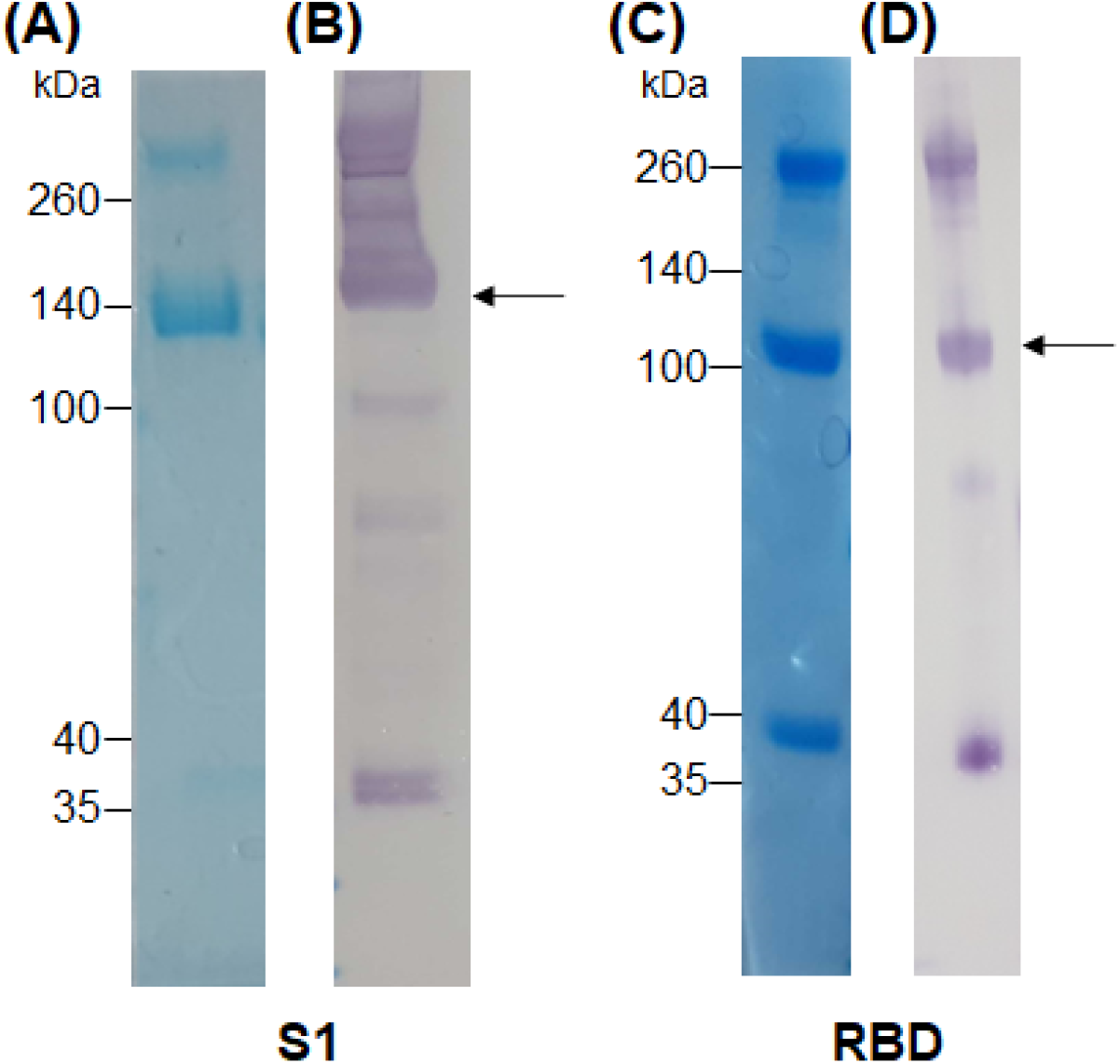
Analysis of plant-expressed SARS-CoV-2 spike antigens after Protein A purification. **(A)** Coomassie-stained SDS-PAGE gel and **(B)** Western blot of S1-rabbit Fc fusion protein (2 µg of concentrated elution fraction). Lines on the left indicate molecular weight marker (Spectra Multicolor Broad range protein ladder) in kDa. The arrow indicates the expected size for recombinant S1 protein (∼140 kDa). **(C)** Coomassie-stained SDS-PAGE gel and **(D)** Western blot of RBD-rabbit Fc fusion protein (5 µg of concentrated elution fraction). Arrows indicate expected size for RBD-rabbit Fc conjugate (∼100 kDa).

### Participant description

Serum samples were collected from 77 volunteers who had previously tested positive for SARS-CoV-2 by PCR. The demographic and clinical characteristics of the participants are summarized in **Table 1**. Just over half the participants were female, and the median age was 39 years. The date of onset of symptoms was not available, but samples were taken a median of 6 weeks after SARS-CoV-2 PCR positivity. The majority of patients (79%) experienced mild or moderate COVID-19 disease. We also included 58 archived plasma samples from HIV-infected and uninfected individuals collected prior to the pandemic (2011-2012) as negative controls for our assay.

**Table 1:** Characteristics of COVID-19 patients (n=77)

|  |  |
| --- | --- |
| Sex female, n (%) | 42 (55) |
| Age (years)<sup>a</sup> | 39 [29-50] |
| Time since positive PCR test (days)<sup>a</sup> | 42 [29-52] |
| Disease severity, n (%)<sup>b</sup> |  |
| *Asymptomatic* | 5 (7) |
| *Mild* | 23 (30) |
| *Moderate* | 38 (49) |
| *Severe* | 2 (3) |

<sup>a</sup> median and interquartile range
<sup>b</sup> not available for n=9 participants

### Plant-produced S1 and RBD proteins are suitable for ELISA detection of SARS-CoV-2 antibodies

In order to test whether plant-produced SARS-CoV-2 antigens were able to detect virus-specific antibodies from infected patients, we screened convalescent sera from 77 volunteers who had recovered from COVID-19. Individuals were tested for reactivity against both S1 and RBD antigens by a standard indirect ELISA based on a published protocol (Stadlbauer et al., 2020). Archived pre-pandemic plasma samples from 58 individuals, including 27 HIV-infected persons, were used to test the background reactivity to SARS-CoV-2 S1 and RBD. The threshold for positivity was set at two standard deviations above the mean optical density (OD) of the pre-pandemic samples.

Of the 77 COVID-19 convalescent serum samples, 51 (66%) tested positive for SARS-CoV-2-specific IgG against S1, and 48 (62%) tested positive against RBD (**Figure 2A & B**). In contrast, only 1/58 pre-pandemic plasma samples showed reactivity above the positivity cutoff. As expected, S1 and RBD IgG OD values correlated strongly (r=0.977; p<0.0001; data not shown). In order to independently validate our results, the same sera were run in a separate laboratory in a blinded manner, using a commercial IgG ELISA based on S1 antigen from Euroimmun. All samples that were positive by the commercial ELISA test for SARS-CoV-2 S1 antibodies were positive in our assay (42/77). We detected nine additional samples that were positive in our assay, two of which had high OD values well above our threshold for positivity, and six that were also positive for RBD-specific IgG. We demonstrated a strong and significant direct correlation for sample OD values between the two assays (r=0.89, p<0.0001; **Figure 2C**). Of note, we found no association between SARS-CoV-2-specific IgG OD values and disease severity or days post PCR positivity (data not shown).

**Figure 2.**
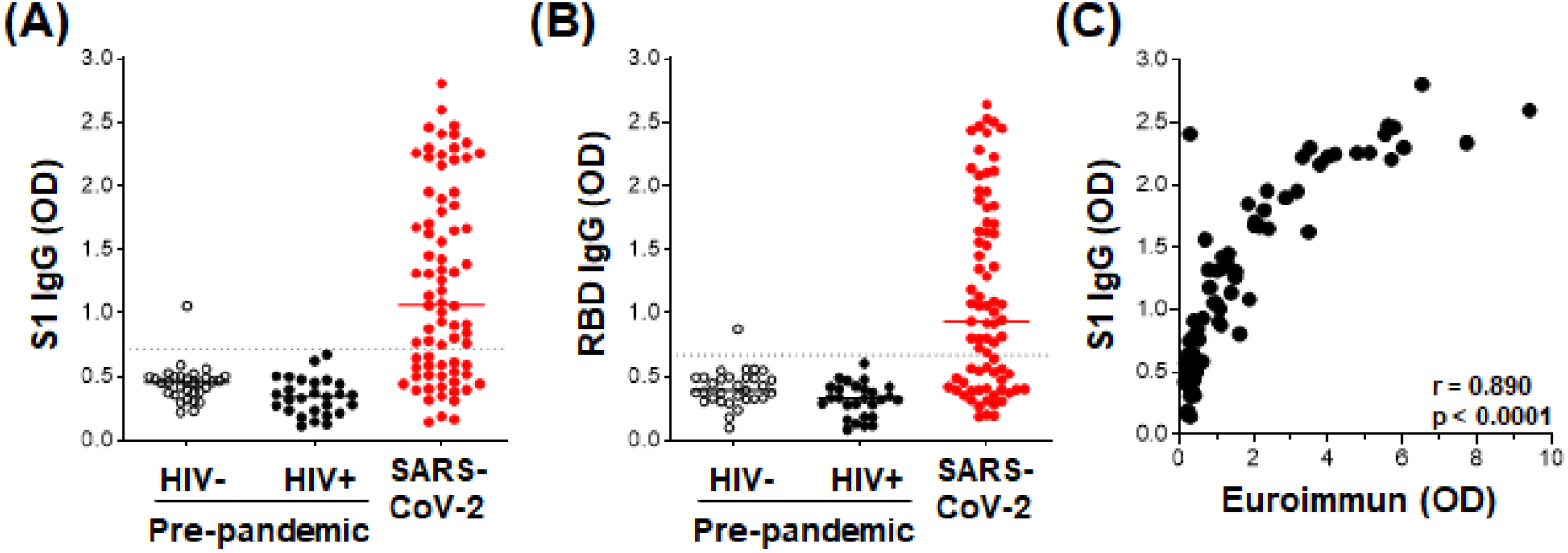
Detection of IgG using plant-expressed SARS-CoV-2 spike antigens in COVID-19 convalescent volunteers and pre-pandemic controls using an in-house ELISA. Reactivity to plant-expressed S1 **(A)** and RBD **(B)** in pre-pandemic samples from HIV-uninfected individuals (n=31), HIV-infected individuals (n=27), and SARS-CoV-2 PCR positive volunteers (n=77). Dotted lines indicate threshold for positivity, calculated as the mean optical density (OD) + 2SD of the pre-pandemic samples. **(C)** Correlation of the OD values for S1-specific IgG in our in-house ELISA and the commercial Euroimmun IgG S1 ELISA kit. Statistical analyses were performed using a non-parametric Spearman rank correlation. Each dot represents one individual.

Thus, our ELISA using plant-produced recombinant viral proteins performed similarly to a highly sensitive and specific commercial SARS-CoV-2 ELISA.

### Determination of immunoglobulin titers and isotypes

We next determined the titers of SARS-CoV-2-specific IgG, IgM and IgA responses in a subset of 20 SARS-CoV-2 convalescent serum samples and 40 pre-pandemic samples. Assays were performed on serially diluted samples (**Figure 3A-F**) to determine endpoint titers and AUC values for quantitative interrogation of the data (**Figure 3G-L**). S1-specific IgG was detected in sera of 15/20 individuals (75%), IgM in 13/20 (65%) and IgA in 12/20 (70%) of individuals (**Figure 3G-I**). The median AUCs of IgG, IgM and IgA were significantly higher in convalescent individuals compared to pre-pandemic (p<0.0001 for all). Results for RBD-specific IgG were similar (**Figure 3J-L**). Interestingly, of the five SARS-CoV-2 convalescent sera that tested S1 IgG negative, three had S1-specific IgM and one had S1-specific IgA. Similarly, of the four samples negative for RBD-specific IgG three were positive for IgM and one was double positive for IgM and IgA. Therefore, SARS-CoV-2 S1-specific antibodies were detected in 19/20 convalescent samples and RBD-specific antibodies in 20/20 samples.

**Figure 3.**
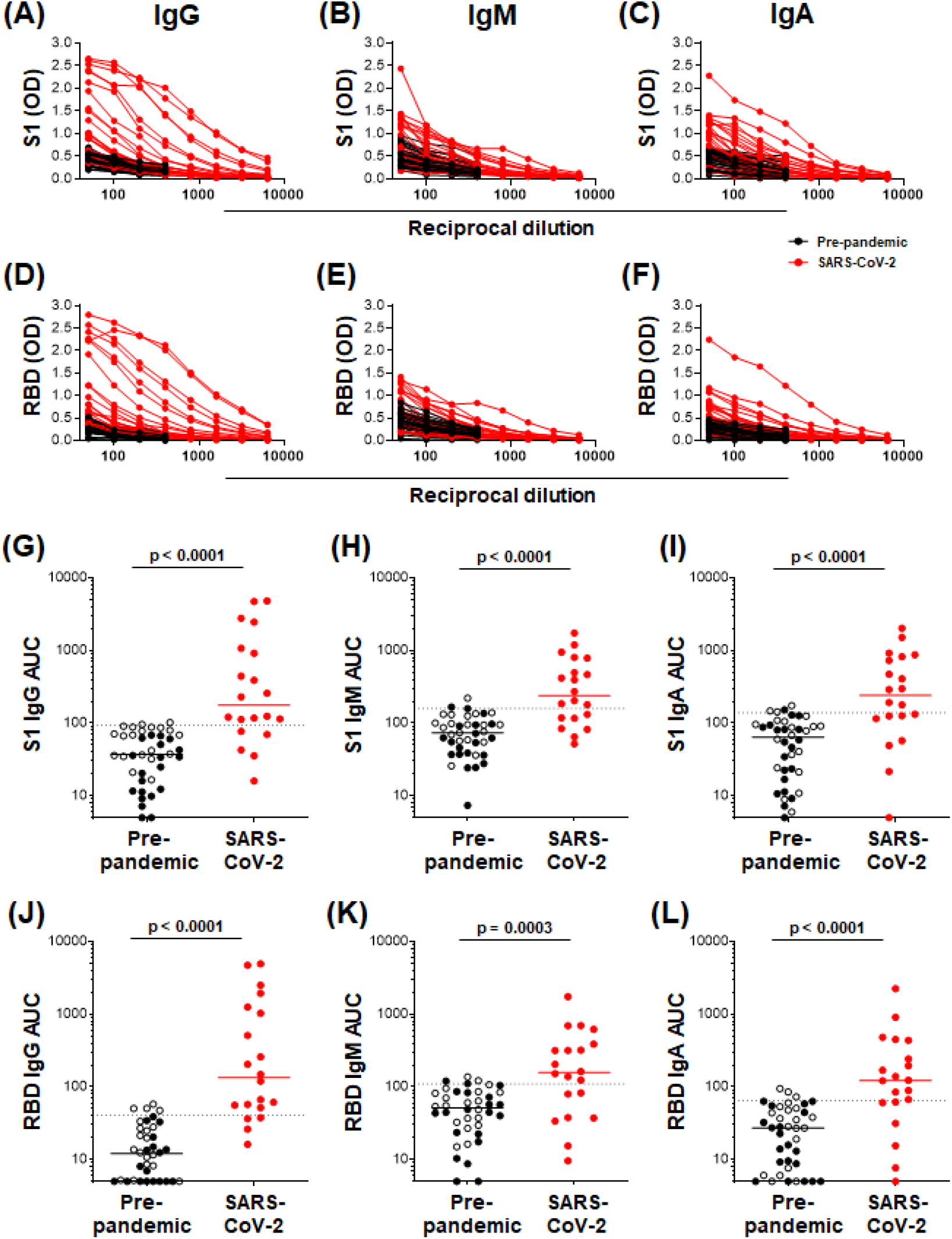
Semi-quantitative detection of S1- and RBD-specific IgG, IgM and IgA. Two– fold dilution series of sera for detection of S1-specific IgG **(A)**, IgM **(B)**, and IgA **(C)** and RBD-specific IgG **(D)**, IgM **(E)** and IgA **(F)**. COVID-19 convalescent volunteers (n=20) are indicated in red, and pre-pandemic controls (n=40) are indicated in black. (**G-I)** and (**J-L)**, Data from the same experiment as in **(A-C)** and **(D-F)**, respectively, but plotted as area under the curve (AUC). Horizontal lines represent median values. Dotted lines indicate the threshold for positivity. Statistical analyses were performed using a Mann-Whitney U test. A p value of <0.05 was considered statistically significant.

Further examination of S1-specific antibody isotypes revealed that approximately one third of individuals were positive for IgG, IgM and IgA (n=7/19), a smaller proportion has both IgG and IgM or IgG and IgA (n=3 and 4, respectively), while some individuals were positive for only IgG (n=1), IgM (n=3) or IgA (n=1) (**Figure 4A**). RBD-specific isotypes gave similar results (**Figure 4B**). There was a significant correlation between S1-specific IgG and IgM (r=0.595, p<0.007; **Figure 4C**) and anti-RBD (r=0.045, p<0.045; data not shown). S1-specific IgG showed a trend towards a correlation with IgA (r=0.423, p=0.07; **Figure 4D**), whilst RBD-specific IgG correlated significantly with IgA (r=0.635, p<0.003; data not shown). There was no correlation between IgM and IgA responses for either S1 or RBD (data not shown).

**Figure 4.**
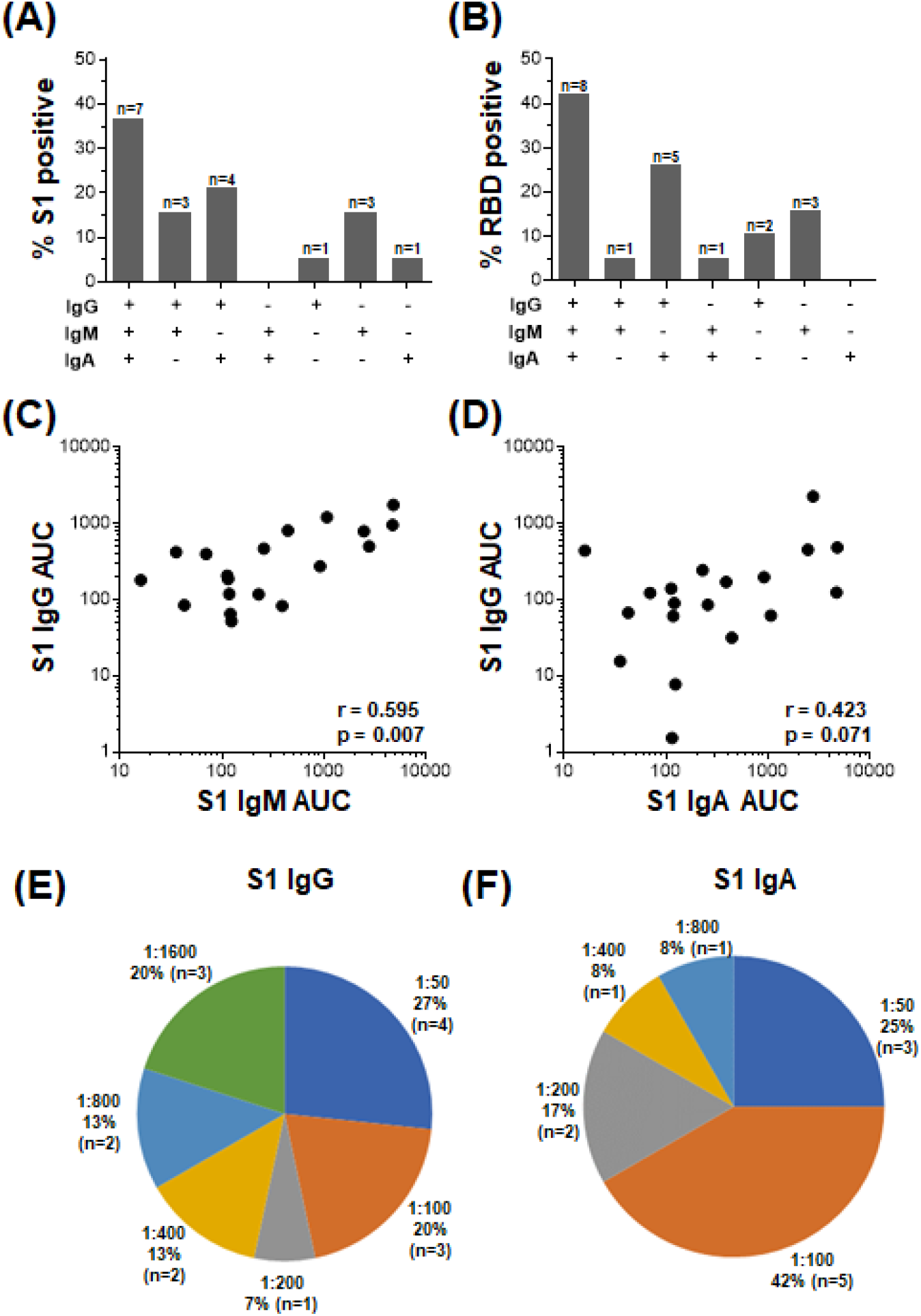
The relationship between IgG, IgM and IgA responses to S1 and RBD SARS-CoV-2 antigens. **A)** Proportions of COVID-19 convalescent volunteers mounting different combinations of IgG, IgM and IgA specific for S1 **(A)** (n=19), and RBD **(B)** (n=20). Relationship between S1-specific IgG and IgM **(C)** and IgG and IgA **(D)**. Statistical analyses were performed using a non-parametric Spearman rank correlation. Proportion of convalescent volunteers with endpoint titers for IgG **(E)** and IgA **(F)** of 1:50, 1:100, 1:200, 1:400, 1:800, 1:1600.

Endpoint titers for S1- and RBD-specific IgG, IgM and IgA were determined. S1-specific IgG endpoint titers in 33% of the samples were high (20% at 1:1600 and 13% at 1:800), 13% were moderate (1:400) and the majority (54%) of samples had low titers (27% at 1:50, 20% at 1:100 and 7% at 1:200) (**Figure 4E**). S1-specific IgA titers were lower than IgG and only 2 individuals have a titer of 1:800 or 1:400 each, and the remaining 84% had low titers (=<1:200; (**Figure 4F**). IgM titers for both S1 and RBD were all low (=<1:100; data not shown). RBD-specific titers for IgG and IgM were similar to those S1, with the exception of two donors who had titers of 1:3200 (data not shown).

### Detection of SARS-CoV-2-specific antibodies in saliva

Given that virus-specific serum antibodies were readily detectable using plant-produced SARS-CoV-2 antigens, we investigated the detection of salivary IgG and IgA using our assay. We compared antibody responses to SARS-CoV-2 antigens in paired saliva and serum from 10 participants. In these preliminary analyses, 1/7 samples that had detectable S1-specific serum IgG also demonstrated S1 IgG positivity in saliva (**Figure 5A**). Additionally, 2/5 IgA+ sera exhibited virus-specific IgA in saliva. An additional IgA+ sample was detected in saliva but absent from the serum (**Figure 5B**). This indicated that IgA was more readily detectable in saliva than IgG. Further analyses to determine robust thresholds for positivity of saliva immunoglobulins will be performed going forward. These preliminary results demonstrate the potential of our ELISA to detect antibodies to SARS-CoV-2 in saliva.

**Figure 5.**
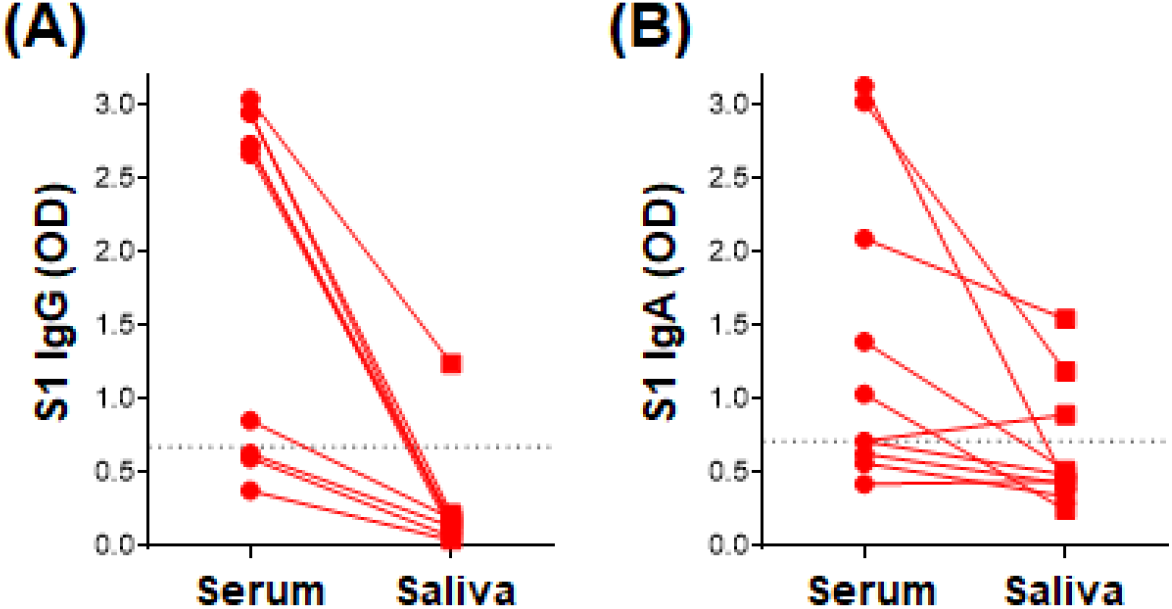
Detection of S1-specific antibodies in saliva. Comparison of paired serum and saliva S1-specific IgG **(A)** and IgA **(B)** (n=10). Dotted lines indicate the positivity threshold for serum.

### Optimization of the ELISA assay

The in-house ELISA diagnostic assay in this study was developed from the published protocol (Stadlbauer et al., 2020). To determine whether we could further improve the robustness and sensitivity of the in-house ELISA assay, we optimized different parameters, including S1 and RBD antigen coating concentration as well as the coating and blocking buffers. Coating concentrations of 1, 2 and 4 □µg/mL S1 and RBD were compared for SARS-CoV-2-specific IgG detection in four SARS-CoV-2 convalescent volunteers and three pre-pandemic samples. Two and 4 µg/ml demonstrated a significantly higher reactivity than 1 µg/ml for both S1 and RBD (**Figure 6A & B;** p=0.0005 and p=0.004, respectively), with little increase in the background (negative control) signal. Coating of ELISA plates with antigen in different coating buffers, namely PBS and bicarbonate buffer, was also assessed (**Figure 6C**). No differences were detected, so PBS was selected for our procedure. A comparison of the blocking buffers PBS with 0.1% Tween-20 (PBS-T), PBS-T with 1% casein and PBS-T with 3% non-fat milk powder was performed (**Figure 6D**). PBS-T with 1% casein was selected based on background signal and positivity trends.

**Figure 6.**
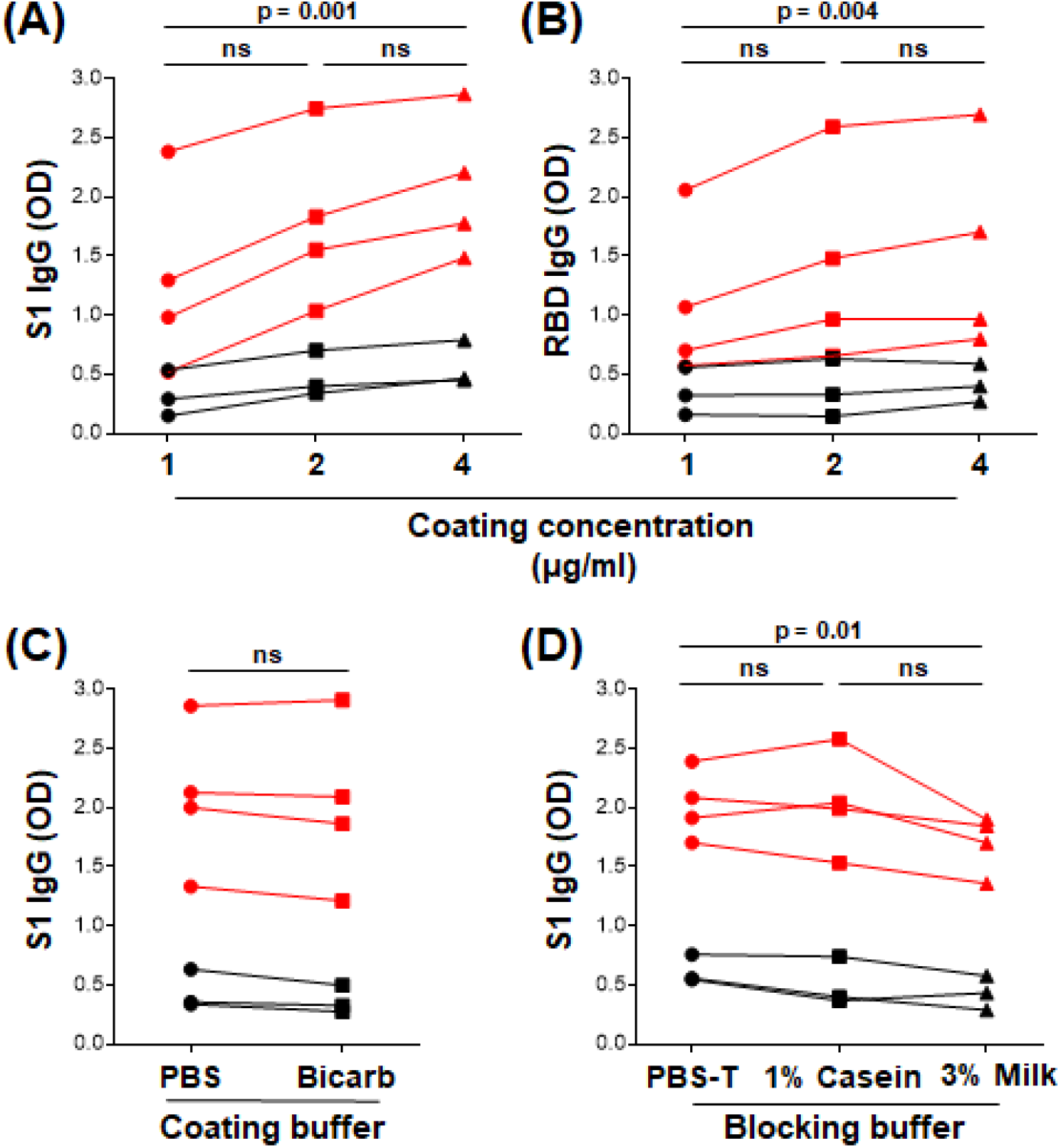
Optimization of ELISA antigen coating concentration, coating buffer and blocking buffer. The effect of antigen coating concentration (1, 2 and 4 μg/ml) was tested for **(A)** S1 and **(B)** RBD, using serum samples from SARS-CoV-2 positive convalescent participants (n=7). Statistical analyses were performed using the Friedman test with Dunn’s test for multiple comparisons. **(C)** Comparison of phosphate buffered saline (PBS) and bicarbonate buffer for coating viral antigens. Statistical analyses were performed using a Wilcoxon matched pair’s test. **(D)** The effect of different blocking solutions. Statistical analysis was performed using the Friedman test with Dunn’s test for multiple comparisons.

## Discussion

There is a critical need for the development of serological tests to detect SARS-CoV-2 antibodies. Population seroprevalence studies to estimate the extent of pandemic spread in communities, and studies defining protective immunity to SARS-CoV-2, all depend on reliable serological tests. In addition, serological assays are required for the development and evaluation of an effective vaccine. Ideally, such tests need to be cost-effective and easy to scale up to be beneficial in low-income settings. In this study, we describe the establishment of an indirect SARS-CoV-2 antibody ELISA using the S1 and RBD antigens of the spike protein of SARS-CoV-2 expressed in *Nicotiana benthamiana*. S protein domains were selected because they are highly immunogenic and the primary target for neutralizing antibodies (Berry et al., 2010; Chen et al., 2020). Using sera from convalescent volunteers with a PCR-confirmed past SARS-CoV-2 infection, we detected SARS-CoV-2-specific IgG, IgA and IgM to viral S1 and RBD. Our results were highly concordant with a widely used, high sensitivity and specificity commercial S1 IgG ELISA kit (Euroimmun).

A range of expression systems exist for the generation of the recombinant proteins required for serological assays. Plant protein expression systems have some advantages over more widely-used mammalian or insect cell systems, as they do not require expensive media or growth conditions (Shanmugaraj et al., 2020). They are also advantageous over bacterial or yeast systems in that they may support post-translational modifications similar to that of mammalian cell lines, and lack contaminating pathogens or endotoxins that pose a problem when purifying desired proteins (Shanmugaraj et al., 2020; Maliga et al., 2004). Lack of correct protein glycosylation and recombinant protein yield are cited as disadvantages to using plants to express protein. However, *Nicotiana benthamiana* is favoured for protein expression due to its rapid generation of biomass, a defective post-transcriptional gene silencing system, and the extensive range of engineering strategies, including glycoengineering, that can be applied along its secretory pathway; all of which may overcome the challenge of low yield (Margolin et al., 2020). Thus, there is great potential to use plant-based expression systems for the rapid generation of serological assay reagents and even vaccines for pandemics, including the current global SARS-CoV-2 pandemic.

Using our ELISA with plant-derived recombinant viral proteins, we detected S1-specific IgG in 66.2% and RBD-specific IgG in 62.3% of individuals who had tested positive for SARS-CoV-2 by PCR in the past. Responses between the two protein fragments were highly correlated, as predicted, and the small difference in reactivity was not unexpected, given the greater number of epitopes in the larger S1 domain. Our sensitivity appears lower than that reported in the literature, with a seroprevalence of 90.1%-100% in individuals confirmed to have been SARS-CoV-2-infected by PCR (Amanat et al., 2020; Beavis et al., 2020; Long et al., 2020; Liu et al., 2020), and a lower seroprevalence (65.8%) in those who were diagnosed <14 days before serological testing (Pollán et al., 2020). However, we obtained highly concordant results between our assay and a validated commercial ELISA. In fact, the reported manufacturer’s sensitivity of the Euroimmun S1-specific IgG ELISA is 94.4%. This suggests that the lack of S1-specific IgG detection from some recovered COVID-19 patients in our cohort is more likely due to low or absent IgG antibody at the time of sampling, rather than a lack of sensitivity in our assay. With regard to specificity, we detected IgG cross-reactivity to SARS-CoV-2 in 1/58 (1.7%) of pre-pandemic plasma samples from a cohort of HIV-infected and uninfected volunteers with latent TB infection, giving a specificity of 98.3%. Cross-reactive antibody responses, while lower in magnitude, have been reported in SARS-CoV-2 unexposed individuals (Khan et al., 2020), and likely result from past infections with common circulating hCoVs. Thus, our assay for SARS-CoV-2-specific IgG performs as well as a widely used commercial kit in terms of sensitivity and specificity, and is suitable for serological studies of humoral responses in the current pandemic.

Several factors may affect antibody detection after SARS-CoV-2 exposure. Timing of sampling is important, with IgM typically arising first, peaking two to three weeks after symptom onset (Long et al., 2020). IgG is typically detected after IgM in serum, peaking at roughly the same time (Huang et al., 2020). However, in SARS-CoV-2 infection, antibodies may not follow this typical pattern of seroconversion (Long et al., 2020; Seow et al., 2020) and seroconversion to a single Ig subclass has been described (Seow et al., 2020). Interestingly, when investigating isotype responses in addition to IgG, we showed that a further 4/20 (20%) donors had S1-specific IgA or IgM. Thus, in our initial screen where 34% of individuals who had previously tested positive for SARS-CoV-2 by PCR had no detectable IgG responses, 20% may have had isotype responses other than IgG. A recent study showed that combined detection of IgG, IgM and IgA increased the overall detection of SARS-CoV-2 antibodies, enabling better identification of infected individuals with low antibody levels (Faustini et al., 2020).

A further factor in detection of antibodies to SARS-CoV-2 is waning of the response over time, which has potentially important consequences for the duration of protective immunity and the risk of reinfection. One study showed a decrease in IgG in half of patients tested, calculating an overall half-life of 36 days for IgG (Ibarrondo et al., 2020). Waning of binding antibody responses to S and RBD has been reported soon after their peak, particularly IgM and IgA antibodies, but IgG responses have shown persistence for greater than 90 days post-illness onset (Seow et al., 2020; Wajnberg et al., 2020). A limitation of our study was that we did not have information on the date of COVID-19 symptom onset in our cohort, limiting our analyses to time post PCR positivity, which did not yield a relationship with antibody positivity or OD value. Additional factors that may also influence antibody generation and kinetics include disease severity, age and comorbidities. We found no relationship between increasing disease severity and antibody positivity or OD value, likely due to the fact that the majority of our study participants had mild to moderate COVID-19.

We determined endpoint titers of binding antibodies to S1 and RBD in a subset of 20 convalescent participants in our cohort. Several studies have demonstrated that binding antibody titers against S correlate with neutralization capacity (Amanat et al., 2020; Okba et al., 2020; Premkumar et al., 2020). A recent study reporting S-specific IgG titers in almost 20 000 patients screened for eligibility as convalescent plasma donors demonstrated that 70% of IgG+ donors had high titers (>1:960) of antibodies (Wajnberg et al., 2020). Importantly, 100% of those with titers >2880 exhibited neutralizing activity (ID_50_ of >1:10). Although we performed our study on a much smaller sample size, we detected titers of S1 or RDB-specific IgG of up to 1:3200. However, the majority of donors (54%) had titers below 1:200, and only a third of samples had high titers >1:800. Unsurprisingly, IgA and IgM titers were lower than IgG titers, and did not exceed 1:800 for IgA and 1:400 for IgM. Further studies characterising antibody titers in recovered COVID-19 patients in our setting are warranted.

Saliva is a non-invasive specimen that can be self-collected and thus represents an attractive sample type for large-scale sampling such as in seroprevalence studies. We demonstrate that our ELISA can detect SARS-CoV-2-specific IgG and IgA not only in serum, but also in saliva. Further optimization and validation will be required to establish the conditions for optimal detection of antibodies in saliva, including the use of pre-pandemic saliva samples. Recent studies have reported the detection of S-specific antibodies in saliva (Faustini et al., 2020; Randad et al., 2020). Faustini et al. (2020) suggested that the use of both serum and saliva samples increased the detection of SARS-CoV-2 antibody responses, reporting substantial discordance between the two sample types. Although preliminary, our results provide the basis for investigating the detection of SARS-CoV-2 antibodies in saliva using antigens expressed in plants.

In conclusion, our study demonstrates that recombinant SARS-CoV-2 proteins produced in plants enable the robust detection of SARS-CoV-2-specific antibodies. One of our aims was to develop a cost-effective serological assay for both large-scale seroepidemiology as well as research studies of SARS-CoV-2 humoral immunity. We achieved this by making use of plants for the production of viral antigens, which has the benefit of rapid scale-up, and sourcing reagents that were available locally and thus available at a lower cost. Our ELISA can be used to evaluate SARS-CoV-2 seroprevalence and describe the kinetics of the humoral immune response in infected individuals. Serological studies in a setting like ours, in South Africa, where comorbidities such as HIV and TB are highly prevalent, are underexplored and can benefit from this assay.

## Data Availability

All data is available and can be obtained by contacting the corresponding author.

## Acknowledgements

We thank Tamlyn Shaw of Cape Bio Pharms. We also thank Markus Sack from Pro-SPR GmbH, Alsdorf, Germany for his assistance in designing the S1 and RBD genes. We thank Muneerah Smith for inactivation and aliquoting of samples. We thank the study participants for providing samples. We are immensely grateful to Florian Krammer and his laboratory at Icahn School of Medicine at Mount Sinai, USA, for rapidly sharing their reagents, protocols and results with the scientific community, and being a model of what our science should be openly accessible and in the service of humankind.

